# Scalable Micro-Credentials for AI Literacy in Healthcare: An AI-Assisted Framework for Expert-Led Education

**DOI:** 10.64898/2026.03.11.26348161

**Authors:** Gabriel Vald, Yusuf Sermet, Ibrahim Demir

**Author notes:** Corresponding Author, Gabriel Vald.

## Abstract

The rapid evolution of clinical guidelines and artificial intelligence has created a velocity gap in medical education, where traditional curricula frequently fail to keep pace with professional practice. Consequently, there is an urgent need for formalized artificial intelligence (AI) micro-credentials to ensure workforce readiness across the entire healthcare ecosystem. Furthermore, existing assessment models for digital credentialing often rely on multiple-choice questions that prioritize pattern recognition over active clinical reasoning, which obscures the underlying logic behind a learner’s choice. This paper introduces a web-based cyberinfrastructure designed to address these challenges by providing an expert-led, AI-assisted platform for the rapid creation and logic-based evaluation of AI micro-credentials. The system architecture utilizes a no-code, node-based visual editor and incorporates local open-source large language models (LLMs) to maintain institutional autonomy and data privacy. A central innovation is the requirement for learners to provide a written rationale for clinical decisions within branching scenarios, which the system evaluates in real-time to assess the underlying thought process for certification. Proof-of-concept modules regarding AI safety in clinical environments and nursing AI workflows were developed to illustrate the platform’s capabilities in managing complex clinical fail-states and recovery pathways. This paper establishes a comprehensive taxonomy of AI competency requirements across hospital roles and proposes a collaborative model to crowdsource an open-access curriculum for scalable AI credentialing in higher education and healthcare.

## 1 Introduction

The rapid integration of artificial intelligence and shifting clinical guidelines has created a significant velocity gap in medical education. Traditional academic curricula often require years to formally update, navigating complex institutional approval processes, while the predictive technologies and evidence-based practices they aim to teach evolve on a scale of months (Masters, 2024). This temporal lag creates a precarious scenario where new graduates and existing medical staff may enter the workforce or continue practice with competencies that are already partially outdated (Bayne & Ross, 2024). This is particularly critical regarding the safe, ethical, and effective application of algorithmic decision-support tools (Ang, 2025). While theoretical educational roadmaps, such as those proposed by Stanford Medicine (2024), provide a high-level framework for curriculum integration, the practical, localized delivery of this content remains a primary hurdle for higher education institutions and teaching hospitals. AI literacy is no longer considered an elective, niche skill restricted to health informaticists; rather, it has emerged as a fundamental, baseline requirement that impacts every single role in the hospital ecosystem to ensure equitable and safe patient care (Crompton and Burke, 2023). To address this paradigm shift, there is an increasing institutional movement toward rapid credentialing and micro-credentials as a primary mechanism for keeping the healthcare workforce current. Competency-based badges, specialized modules, and targeted digital certificates provide a flexible, modular alternative to traditional degree structures, allowing for the rapid distribution of specialized knowledge as new clinical challenges and software systems emerge (Kovilpillai et al., 2024; Rullyana et al., 2025).

However, the transition to digital, automated, and micro-credentialed education has highlighted a persistent, systemic limitation in student assessment and credential validation. Most current learning management systems and credentialing platforms rely heavily on multiple-choice questions (MCQs), which frequently prioritize superficial pattern recognition over active clinical recall and deep critical reasoning (van Wijk et al., 2025). Research indicates that MCQs often suffer from a pronounced cueing effect, where the mere presence of the correct answer among a list of distractors allows a student to identify the right choice through elimination without necessarily understanding the underlying clinical logic (Lertsakulbunlue & Kantiwong, 2024). This dynamic creates a pedagogical black box in digital assessment: educators and credentialing bodies can verify what a student chose, but they cannot verify the why behind the decision. For an AI micro-credential to be meaningful, reliable, and safe, it must actively evaluate a learner’s ability to critically oversee, critique, and contextualize algorithmic outputs (Zawacki-Richter et al., 2019). Eye-tracking experiments have demonstrated that very short answer questions (VSAQs) and rationale-based assessments require significantly higher cognitive engagement and deeper information processing compared to the passive pattern recognition utilized in MCQs. These reasoning-based assessments provide a substantially higher discrimination index in knowledge evaluation, effectively separating learners who truly understand the clinical context from those relying on test-taking strategies (Potter & McLachlan, 2025; Carneiro Queiroz et al., 2026).

Branching narratives and high-fidelity digital simulations offer a potent pedagogical solution to these assessment gaps by allowing learners to navigate complex, non-linear clinical pathways. Unlike static, linear case studies, error-based branching scenarios force students to actively manage the downstream consequences of their decisions within a psychologically safe environment (Stathakarou, 2026). This mimics the inherent uncertainty of real-world clinical practice and promotes deep critical reflection without risking patient safety (Marcu et al., 2022). While these simulations are highly predictive of clinical competence, they are rarely implemented at scale due to immense design complexity (Gonçalves et al., 2026). These simulations have proven highly effective for multidisciplinary training and high-stakes clinical environments, such as the neonatal intensive care unit (NICU), where procedural foresight and the ability to interpret complex data streams are paramount to preventing adverse events (Carvalho et al., 2025).

Despite their clear, documented pedagogical value, the widespread creation and deployment of such simulations remains technically prohibitive for most educational institutions (Bozkurt et al., 2023). Subject matter experts, the frontline clinicians who possess the nuanced knowledge required for realistic scenarios, often face insurmountable barriers when attempting to author these high-fidelity materials. The steep learning curve of traditional simulation software, coupled with a chronic lack of technical support staff in academic medicine, frequently prevents clinicians from translating their expertise into scalable digital formats (Huertas-Zurriaga et al., 2026; O’Doherty et al., 2018). Furthermore, institutional cultures may fail to recognize these complex authoring efforts as formal academic scholarship, disincentivizing participation. There is a critical, unmet requirement for low-code or no-code cyberinfrastructures that allow medical professionals to create credentialing content rapidly, removing technical friction (Ukeje, 2023; Stenalt & Mathiasen, 2024). By strategically utilizing large language models (LLMs) as localized teaching aides, the cognitive burden of drafting, structuring, and mapping these complex narratives can be significantly reduced, allowing for the technical feasibility of high-fidelity simulations with a notable reduction in content creation time (Nouraei et al., 2025; Salim et al., 2025).

This paper introduces a novel, web-based cyberinfrastructure designed specifically to bridge these educational and technical gaps by providing an expert-led, AI-assisted platform tailored for medical education and AI micro-credentialing. The system focuses on three core technological and pedagogical innovations: (i) an intuitive, node-based visual editor designed for rapid, frictionless scenario creation by non-technical clinical experts; (ii) a strict pedagogical requirement for learners to provide a written rationale for their clinical choices at critical decision nodes; and (iii) a secure, localized LLM-based feedback loop that qualitatively evaluates the logic of those rationales in real-time. This approach moves institutional credentialing beyond basic, binary AI awareness toward a rigorous framework of algorithmic critique, where learners are actively taught and assessed on their ability to safely oversee and correct AI outputs (Burneo-Arteaga et al., 2025). To validate this infrastructure, proof-of-concept modules were developed and evaluated with clinical educators. This paper therefore aims to serve as an open invitation for international collaboration; by outlining a comprehensive taxonomy of AI competencies required across the hospital ecosystem, we propose a community-driven model to crowdsource an open-access, role-specific curriculum for scalable AI credentialing.

## 2 Methods

### 2.1 A Taxonomy of AI Competency in the Hospital Ecosystem

The integration of artificial intelligence into clinical, diagnostic, and administrative workflows necessitates a fundamental paradigm shift in how higher education and healthcare institutions approach professional development and credentialing. It is pedagogically and operationally insufficient to issue a universal, one-size-fits-all “AI in Healthcare” certificate. The required competencies, ethical considerations, cognitive loads, and safety protocols vary drastically depending on a stakeholder’s specific role and scope of practice within the healthcare environment. For instance, an interventional radiologist’s interaction with an AI-driven predictive imaging tool requires a fundamentally different skill set, focusing on diagnostic momentum and algorithmic opacity, than a clinical documentation improvement (CDI) specialist utilizing automated coding software, or an intensive care nurse managing predictive deterioration alerts and mitigating alarm fatigue.

Consequently, to establish a robust, scientifically grounded framework for official institutional AI credentialing, it is imperative to map the entire hospital ecosystem and identify the unique professional intersections where each specialty encounters algorithmic decision-making. To systematically address these varied educational requirements, we have developed a comprehensive taxonomy of AI competency targets encompassing the operational and clinical specialties within a modern healthcare institution. As detailed in Table 1, this taxonomy categorizes stakeholders into distinct functional domains, serving as the foundational blueprint for a targeted micro-credentialing curriculum.

**Table 1.** Comprehensive Stakeholder Taxonomy for Role-Specific AI Credentialing.

| **Functional Domain** | **Target Professions and Specialties** |
| --- | --- |
| **Physicians & Medical Providers** | Anesthesiology, Allergy & Immunology, Cardiology (General, Interventional, Electrophysiology, Heart Failure), Critical Care/Intensive Care, Dermatology, Emergency Medicine, Endocrinology, Family Medicine, Gastroenterology, Geriatric Medicine, Hematology/Oncology, Infectious Diseases, Internal Medicine, Medical Genetics, Nephrology, Neurology, Neurosurgery, OB/GYN, Ophthalmology, Orthopedic Surgery, Otolaryngology, Palliative Medicine, Pathology, Pediatrics, PM&R/Physiatry, Plastic Surgery, Preventive Medicine, Psychiatry, Pulmonology, Radiation Oncology, Rheumatology, Sleep Medicine, Sports Medicine, Thoracic Surgery, Urology, Vascular Surgery. |
| **Dental & Oral Health** | General Dentist, Oral & Maxillofacial Surgeon, Orthodontist, Endodontist, Periodontist, Prosthodontist, Pediatric Dentist, Dental Hygienist, Dental Assistant. |
| **Advanced Practice Providers** | Nurse Practitioner (Family, Acute, Psychiatric), Physician Assistant / Associate, Certified Nurse Midwife, Anesthesiologist Assistant, Clinical Nurse Specialist. |
| **Nursing Workforce** | Registered Nurse (Inpatient, Outpatient, ED, OR, ICU, Oncology), LPN / LVN, Certified Nursing Assistant, Nurse Anesthetist, Nurse Informaticist, Nurse Educator, Home Health / Visiting Nurse, Occupational Health Nurse, School Nurse, Public Health Nurse, Case Manager, Patient Navigator. |
| **Allied Health Professionals** | Physical Therapist, Occupational Therapist, Speech-Language Pathologist, Respiratory Therapist, Radiologic / X-ray Technologist, Sonographer, MRI Technologist, Nuclear Medicine Technologist, Radiation Therapist, Surgical Technologist, Perfusionist, Electroneurodiagnostic Technologist, Ophthalmic Technician, Optometrist, Audiologist, Dietitian, Clinical Exercise Physiologist, Prosthetist & Orthotist, Athletic Trainer. |
| **Laboratory & Diagnostics** | Medical Laboratory Scientist / Technologist, Clinical Laboratory Technician, Phlebotomist, Histotechnologist, Cytotechnologist, Microbiology / Molecular Diagnostics Technologist, Blood Bank / Transfusion Specialist. |
| **Imaging & Radiology** | Radiologist (Diagnostic, Interventional), Radiology Technologist, PACS Administrator, Teleradiology Clinicians. |
| **Behavioral Health & Support** | Psychologist (Clinical / Counseling), Licensed Clinical Social Worker, Mental Health / Licensed Professional Counselor, Marriage & Family Therapist, Substance Use Disorder Counselor, Behavioral Health Nurse, Mental Health Case Manager. |
| **Health Info, Coding & Revenue** | Health Information Manager, Medical Coder, Clinical Documentation Improvement Specialist, Billing Specialist / Revenue Cycle Analyst, Medical Transcriptionist / Clinical Scribe, Release of Information Specialist. |
| **Quality, Safety & Compliance** | Quality Improvement Specialist, Patient Safety Officer, Compliance Officer, Risk Manager, Medical-Legal Consultant, Privacy / Data Protection Officer. |
| **Administrative & Operations** | Medical Office Manager, Receptionist / Scheduling Coordinator, Patient Access Representative, Medical Secretary, Patient Experience Manager, Complaint / Grievance Specialist. |

Our primary objective with this cyberinfrastructure is to establish a targeted, foundational curriculum for each specialty outlined in this taxonomy, combining these role-specific scenarios with location-based regulatory requirements and distinct institutional standards. While the technological infrastructure has been successfully developed to host, adapt, and deliver these customized educational pathways, generating the requisite high-fidelity clinical scenarios for every specialized area requires immense, localized domain expertise. We view this credentialing system not as a static software product, but as a growing, collaborative mechanism designed to provide official credentialing for AI capacity across the medical sector. To achieve this unprecedented scale, we are utilizing a GitHub repository to foster open academic conversation and invite the global medical education community to participate. By creating issues, mapping logical pathways, and proposing clinical narratives through this collaborative channel, subject matter experts can partner with educational technologists to deploy these specialized curricula, in order to make sure that the resulting micro-credentials reflect the highest standards of frontline practice.

### 2.2 Cyberinfrastructure Architecture for Rapid Credential Development

The architecture of the platform is engineered as a modular, multi-tier cyberinfrastructure specifically optimized to support the demands of institutional credentialing, rapid content authoring by non-technical faculty, and a high-fidelity trainee experience (Figure 1). The system utilizes a multi-page web framework to maintain a strict, functional separation between public access, student engagement, and administrative oversight. This structural organization is essential for ensuring that educational delivery remains distinct from content management, reducing cognitive load for clinical educators and providing a highly secure environment for institutional credentialing data (Hamad et al., 2024). Initial entry occurs through a landing page that facilitates secure user onboarding via OAuth-based authentication, allowing for standardized access control and data protection across different institutional environments.

**Figure 1.**
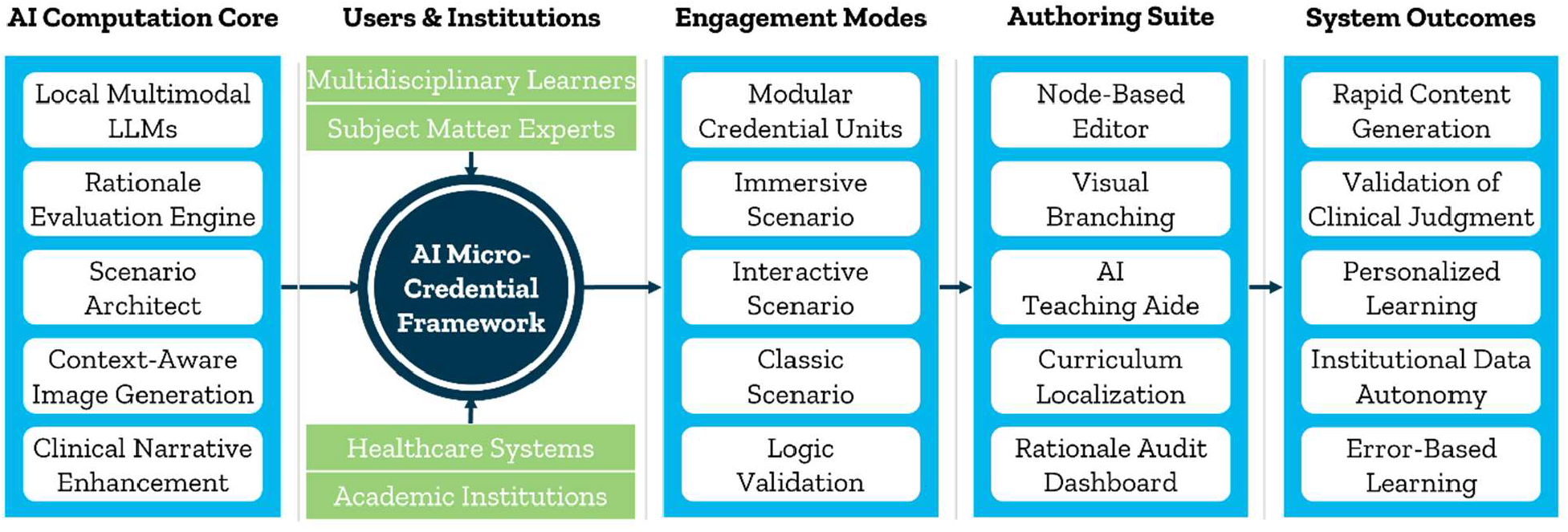
Cyberinfrastructure Architecture mapping the separation of public, trainee, and administrative portals.

The overall technical stack is built on a foundation of React and Node.js for the responsive frontend interface, while the heavy computational logic and data management layers are handled by a Python-based backend. Data persistence, a critical factor for credential auditing and compliance, is managed via a relational PostgreSQL database. This database is specifically structured to handle the relational complexity of non-linear branching narratives, storing course hierarchies, scenario maps, and, most importantly, detailed student audit logs that capture every rationale provided during an assessment.

### 2.3 Node-Based Scenario Architect

To address the technical friction that historically prevents subject matter experts from authoring digital simulations, a critical component of the system is the visual scenario editor, integrated directly into the administrative portal utilizing React Flow. This tool enables clinical educators to map complex clinical narratives and AI competency pathways as a series of visual, interconnected nodes, representing decision points, information updates, and clinical outcomes. To further expedite this process, instructors are provided with a dedicated LLM workshop interface that functions as a conversational aide for drafting modular units. When moving to the visual editor, experts can build narratives from scratch, utilize pre-defined case study templates that provide a structural head start, or use automated tools to generate entire node structures based on uploaded course documents (Sajja et al., 2023).

As shown in Figure 2, this visual mapping approach allows experts to design complex, non-linear educational pathways without requiring engagement with underlying code or scripting languages. Instructors can effortlessly build (i) recovery loops that redirect learners to safe pathways after a mistake, encouraging experiential learning, and (ii) terminal fail-states that clearly illustrate the severe clinical consequences of incorrect decisions or over-reliance on automated systems.

**Figure 2.**
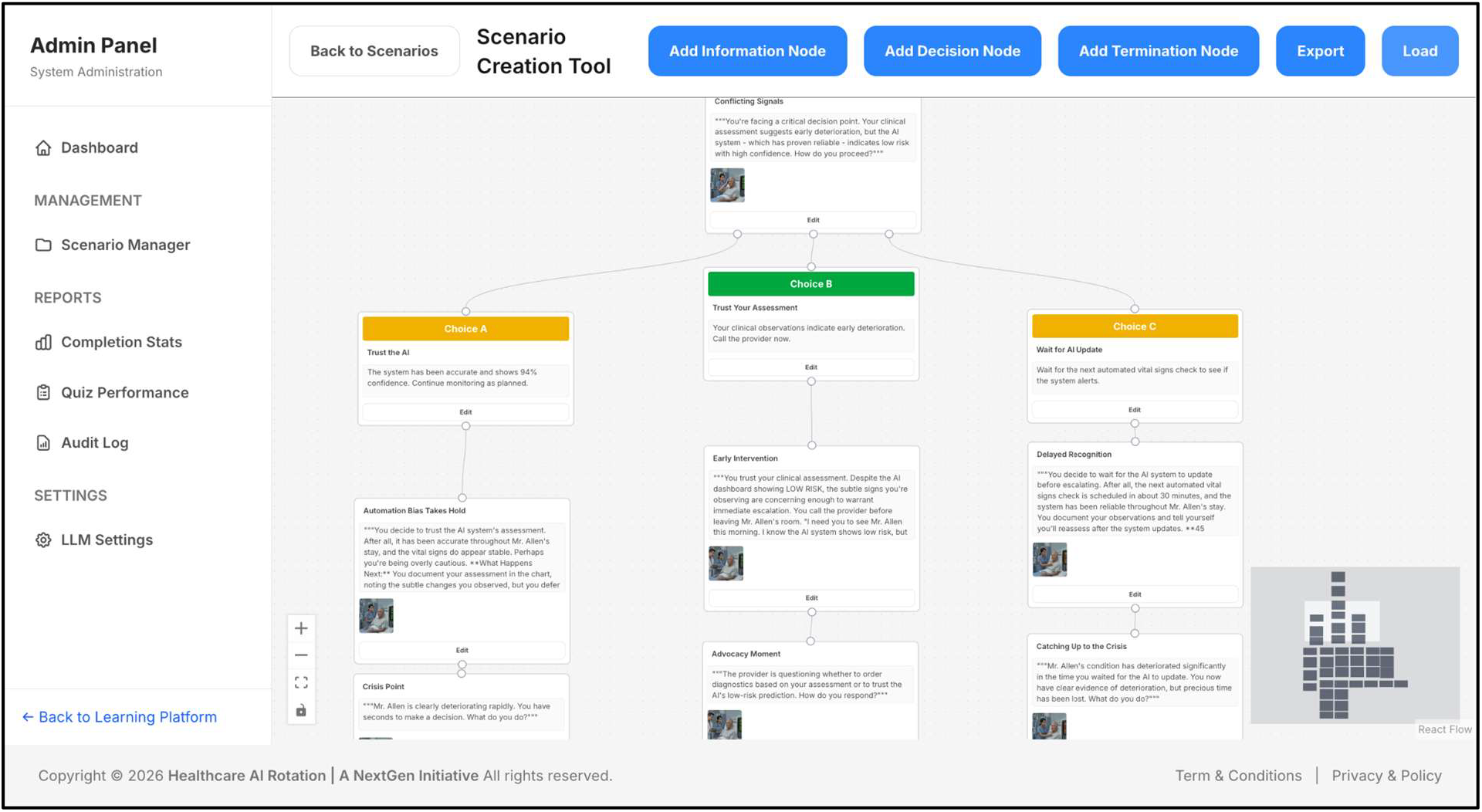
Visual Interface of the Node-Based Scenario Editor demonstrating non-linear pathway mapping.

### 2.4 Local AI Infrastructure and Institutional Autonomy

A defining characteristic of the platform’s architecture, and a non-negotiable requirement for many hospital IT governance boards, is the deployment of local, open-source LLMs for both rationale assessment and content authoring assistance. The system bypasses external, proprietary APIs in favor of locally hosted models such as Llama, Qwen, and DeepSeek. This local deployment strategy ensures total institutional autonomy and data sovereignty (An et al., 2025). It safeguards against the inadvertent exposure of simulated patient health information (PHI) or proprietary institutional protocols to third-party AI vendors (Sajja et al., 2026). Furthermore, by avoiding reliance on commercial APIs, the credentialing platform mitigates the risks associated with vendor lock-in, unexpected pricing surges, or sudden deprecation of model functionalities. This localized infrastructure ensures the system is technologically prepared to rapidly incorporate custom, fine-tuned medical LLMs as they become available, allowing the credentialing infrastructure to scale securely alongside the needs of the medical education community.

### 2.5 Adaptive Delivery Modes and Presentation Architecture

To accommodate the diverse cognitive styles and technical constraints of a sprawling healthcare workforce, the trainee portal utilizes a decoupled content-presentation architecture. This design separates the underlying clinical data and scenario logic from the visual interface, allowing the platform to render the exact same clinical narrative through fundamentally different visual lenses. As detailed in Table 2, learners engaging with an AI micro-credential may choose from three distinct engagement modes based on their learning preferences or specific pedagogical requirements.

**Table 2.** Scenario Delivery Methods and Pedagogical Strengths.

| Delivery Mode | Presentation Style | Pedagogical Strength | Technological Novelty & AI Integration |
| --- | --- | --- | --- |
| Classic Mode | Uniform, accessible presentation resembling a traditional digital case study. Text and static images are embedded within a linear reading flow. | **Efficiency & Comprehension:** Ideal for rapid review or professional settings with limited bandwidth. Minimizes cognitive friction for learners accustomed to traditional textbooks. | **Dynamic Text Assembly:** Automatically compiles non-linear branching node data into a cohesive, easily readable document format in real-time. |
| Interactive Mode | Dialogue-based interface simulating a text-message or chat environment. A virtual AI agent acts as the scenario deliverer, guiding the user. | **Socratic Engagement:** Best for probing the depth of user understanding. The conversational format feels natural and encourages learners to process information through active dialogue. | **Agent-Led Delivery:** Utilizes LLMs to function as an active instructional guide, dynamically delivering narrative prompts and managing conversation flow (Ding et al., 2023). |
| Immersive Mode | Visual-first presentation utilizing high-fidelity imagery paired with a scrolling typewriter text effect at the base of the screen. | **Sustained Focus:** The Rapid Serial Visual Presentation (RSVP) effect regulates reading pace to prevent cognitive overload and maintain deep engagement during complex clinical narratives. | **Context-Aware Visuals:** The AI Content Guide assists in generating custom, contextually relevant imagery for each decision node on the fly. |

The system maintains continuous state-persistence across these modes, allowing a learner to switch between a Classic textual review and an Interactive Socratic dialogue mid-scenario without losing assessment progress. This architectural flexibility ensures that the core educational objectives of the credential remain consistent, regardless of how the learner chooses to interface with the material.

### 2.6 Logic-Based Assessment for Credential Validation

To confidently issue an official, institution-backed AI micro-credential, universities and healthcare systems must be able to scientifically certify that a practitioner can safely oversee, critique, and correct algorithmic outputs in high-stakes environments. However, traditional digital credentialing platforms inherently rely on assessment models that fail to capture this necessary depth of clinical judgment. To systematically address the “cueing effect” prevalent in standard multiple-choice testing, where learners frequently rely on the process of elimination rather than generating true clinical knowledge (Lertsakulbunlue & Kantiwong, 2024), this cyberinfrastructure implements a mandatory, open-ended rationale assessment at every critical decision point.

When a learner encounters a branching decision node within an AI credentialing scenario, the system explicitly does not immediately reveal the outcome or correct pathway associated with their selected option. Instead, the learner is confronted with a cognitive hard-stop: they must provide a written, text-based justification for their clinical decision in an open-ended input field.

This interaction (Figure 3) forces the learner to actively synthesize their clinical knowledge, recall relevant institutional protocols, and explicitly justify their reasoning before the simulation will advance. Once the rationale is submitted, the localized backend LLM evaluates the learner’s input by triangulating three distinct data points: (i) the specific clinical option selected, (ii) the ground-truth pedagogical correctness defined by the curriculum author, and (iii) the qualitative nuance of the learner’s written logic.

**Figure 3.**
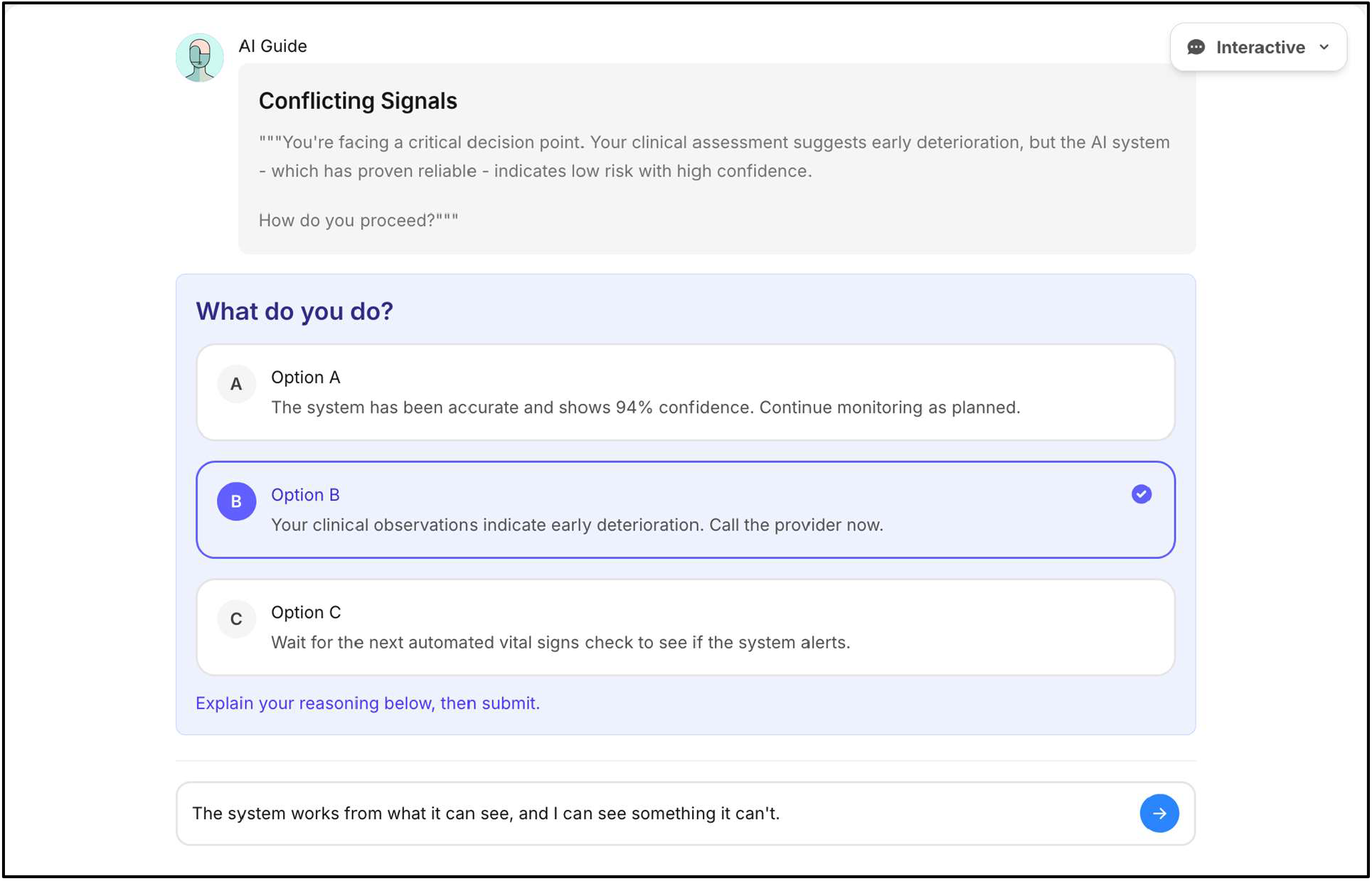
User Interface for Mandatory Rationale Submission requiring text-based justification prior to advancing the scenario.

The system then utilizes this triangulation to generate real-time, conversational, and highly specific feedback that addresses the underlying quality of the learner’s reasoning (Sajja et al., 2024). This feedback mechanism is particularly vital for AI micro-credentialing because it allows the system to identify instances where a student reaches a correct clinical action through deeply flawed or highly dangerous logic. For example, if a learner correctly chooses to administer a specific medication, but their written rationale reveals they did so solely because “the AI suggested it,” rather than citing the patient’s bedside vitals or lab results, the system will flag this as a critical failure of algorithmic oversight. The LLM then provides immediate, targeted remediation that corrects the underlying automation bias. This real-time feedback loop allows learners to internalize the ‘why’ behind clinical best practices, moving the credentialing assessment away from a binary correct-or-incorrect model and establishing a standard for qualitative algorithmic critique.

To support the validation of these credentials, the administrative dashboard provides curriculum directors and compliance officers with granular, transparent visibility into learner performance. Rather than viewing opaque, binary scores, administrators can audit the individual, time-stamped rationale entries for every learner across a given scenario. This data-driven approach supports strict institutional compliance and provides a highly nuanced, verifiable foundation for certifying that future clinicians possess the reasoning skills required to safely navigate an AI-augmented healthcare environment.

## 3 Results

The functional utility of the platform as a foundational cyberinfrastructure for medical AI credentialing was empirically validated through the deployment of specialized proof-of-concept modules. These modules illustrate the effectiveness of the rapid authoring workflow, the adaptive logic of the branching scenarios, and the platform’s capacity to handle clinical decision points, error-based fail-states, and recovery pathways.

### 3.1 System Demonstration: AI Safety Case Study

A foundational module titled “AI Safety in Clinical Environments” was developed to address a ubiquitous gap in health informatics: the psychological tendency of clinicians to over-rely on automated decision-support systems. This module targets core learning objectives related to (i) algorithmic bias, (ii) the preservation of independent clinical judgment, and (iii) the risks associated with data latency in electronic health records. To illustrate these theoretical concepts practically, a clinical scenario was designed wherein a learner must manage a patient showing subtle, physical signs of deterioration while a highly reliable AI predictive system paradoxically indicates a low risk status with high statistical confidence. It is important to note that this foundational unit was authored by platform developers, who possess expertise in health informatics but lack formal clinical training. This distinction underscores the platform’s primary value proposition: providing a no-code authoring environment so that actual frontline clinicians can easily replace this proof-of-concept with nuanced, real-world medical guidelines.

As seen in Figure 4, learners navigate this tension through the Immersive Mode viewer. In Figure 4a, the learner is presented with a critical decision point where the patient’s physical symptoms of deterioration directly contradict a confident, “low risk” algorithmic assessment. In Figure 4b, a downstream fail-state illustrating the severe clinical consequences of automation bias, demonstrating the adverse patient outcome if the practitioner improperly defers to the AI system. The underlying logic map of this scenario, constructed using the administrative visual node editor, is complex. It encompasses standard correct pathways (green nodes), nuanced recovery pathways (yellow nodes), and terminal fail-states (red nodes).

**Figure 4.**
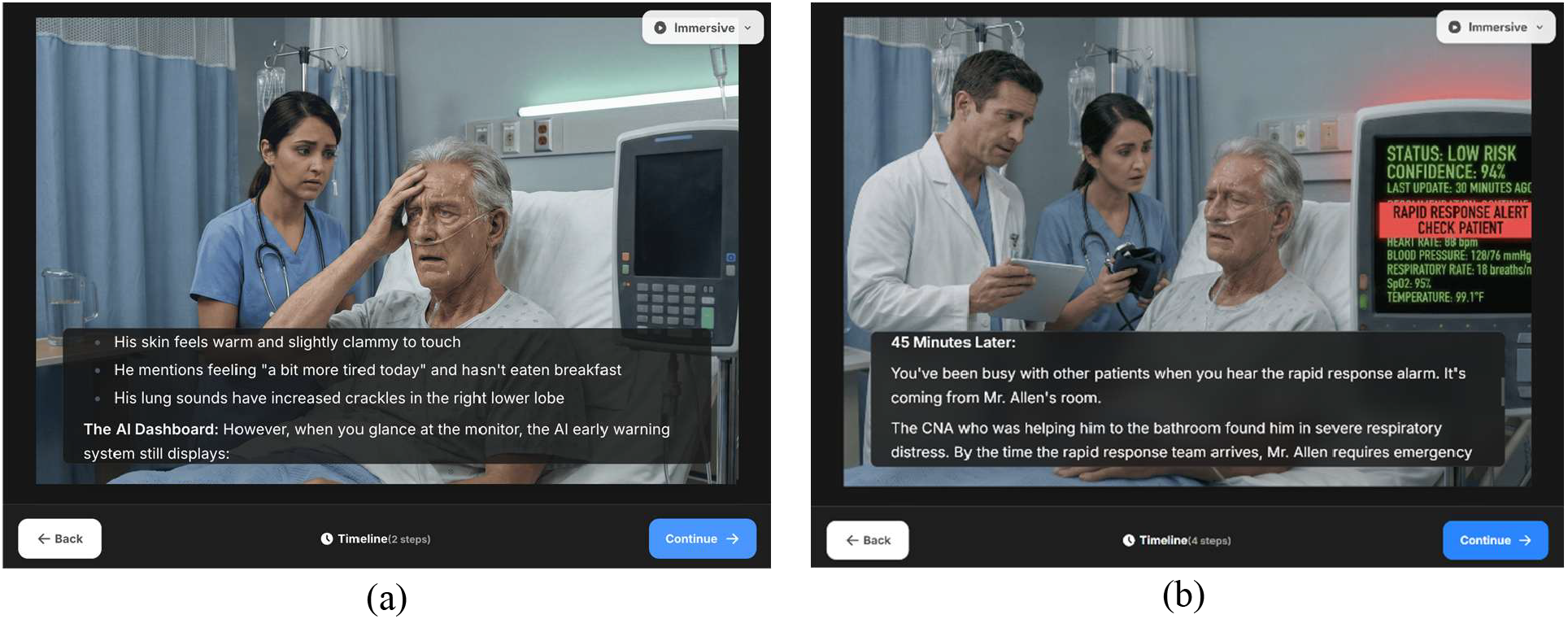
Student Interface for the Immersive Scenario Viewer displaying a high-fidelity clinical narrative.

In the configuration detailed in Figure 5, learners who make a premature or incorrect decision regarding the AI’s output are directed to yellow recovery nodes. These pedagogical detours allow the learner to witness the immediate, negative downstream effects of their choice, such as a sudden drop in the patient’s oxygen saturation, while still providing an educational opportunity to navigate back to a safer clinical pathway. However, if a learner persists in a dangerous line of reasoning across two consecutive decision points, the scenario permanently terminates in a red fail-state, triggering a comprehensive remediation text on the specific dangers of automation bias. This pedagogical structure ensures that learners are not merely punished for mistakes, but are instead guided through a clear, experiential explanation of the clinical risks associated with their flawed reasoning.

**Figure 5.**
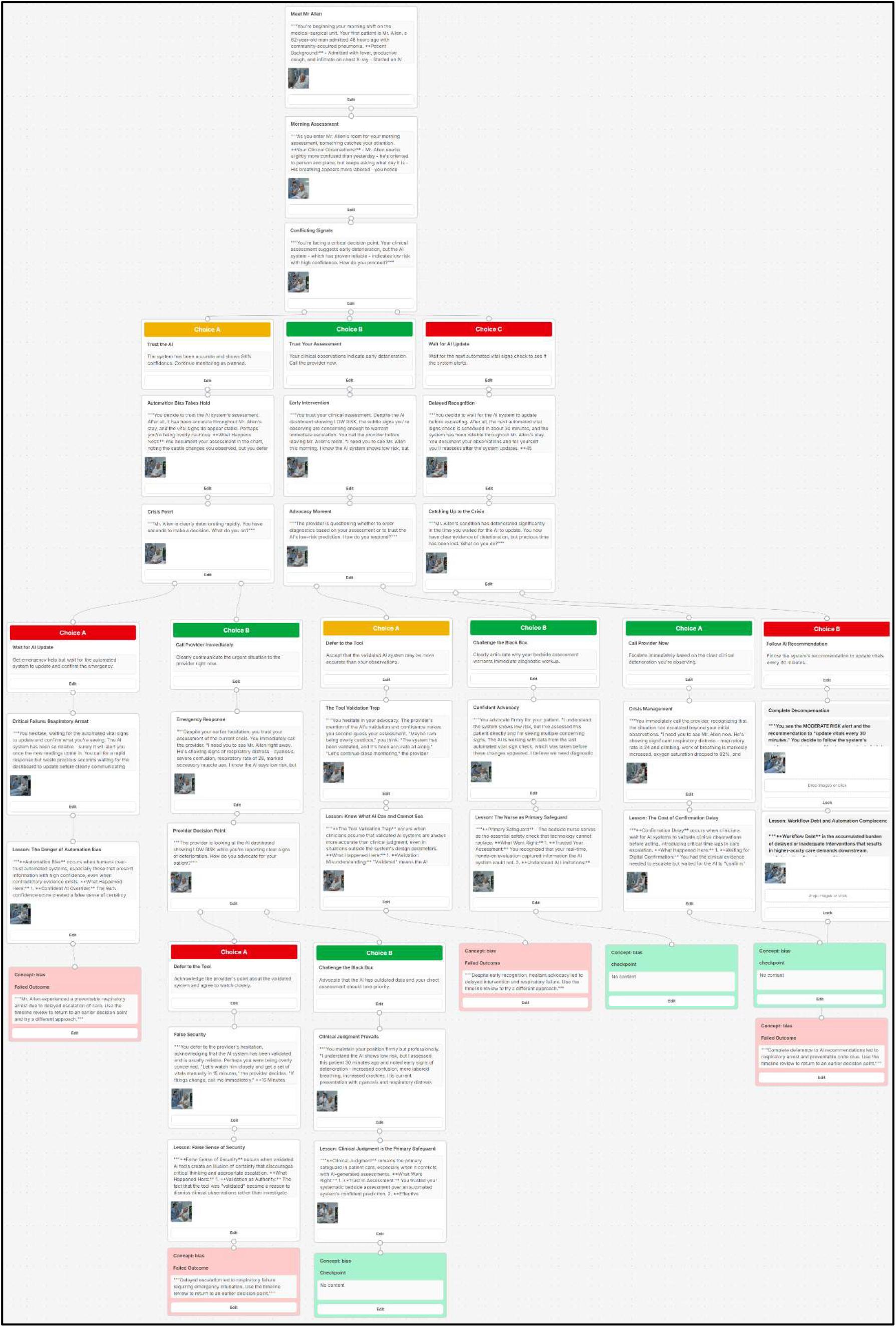
Logic Map of AI Safety Scenario in the Node-Based Editor with recovery and failure pathways.

### 3.2 Validation of the Logic-Based Feedback Loop

The pedagogical effectiveness of the rationale assessment was validated through a series of student-interaction simulations. In these controlled tests, the localized LLM grader successfully identified the nuanced quality of learner justifications across vastly different clinical scenarios.

As illustrated in Figure 6, one of the system’s benefits lies in its ability to provide constructive remediation even when a learner selects the nominally correct clinical choice but provides incomplete logic. Conversely, when a learner demonstrates highly sound clinical judgment, such as explicitly choosing to trust their physical bedside assessment over a confident, yet lagging, AI signal, the grader provides immediate positive reinforcement that actively validates their critical thinking approach.

**Figure 6.**
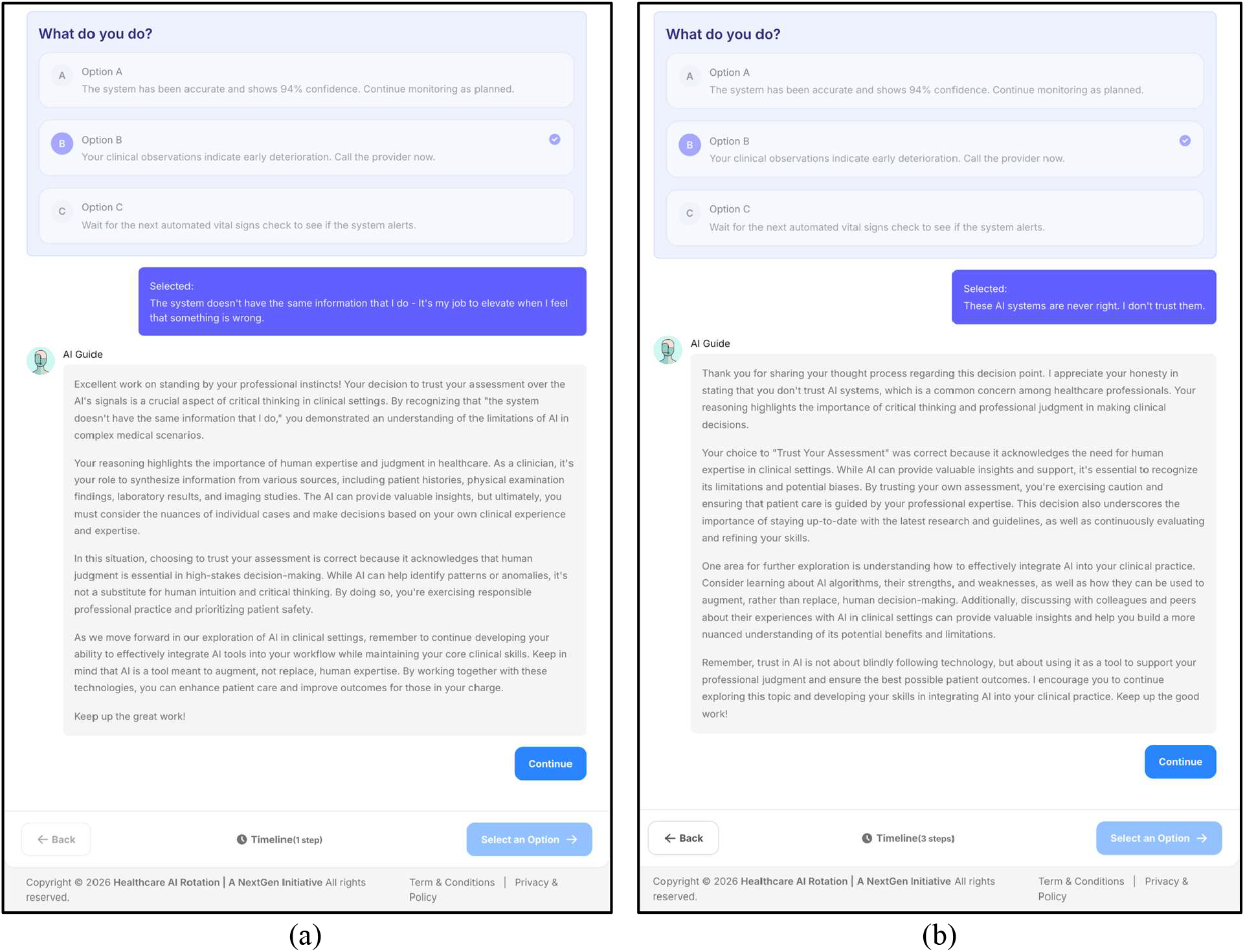
System feedback demonstrating (a) positive reinforcement for sound logic and (b) targeted remediation for flawed logic.

### 3.3 Expanding the Curriculum: Department-Specific Scenario Mapping

To demonstrate the platform’s capacity to scale across the extensive taxonomy outlined in Section 2, a series of departmental scenario frameworks were mapped. These frameworks, detailed in Table 3, illustrate how abstract AI credentialing concepts can be translated into highly specific, relevant simulations for different medical specialties.

**Table 3.**
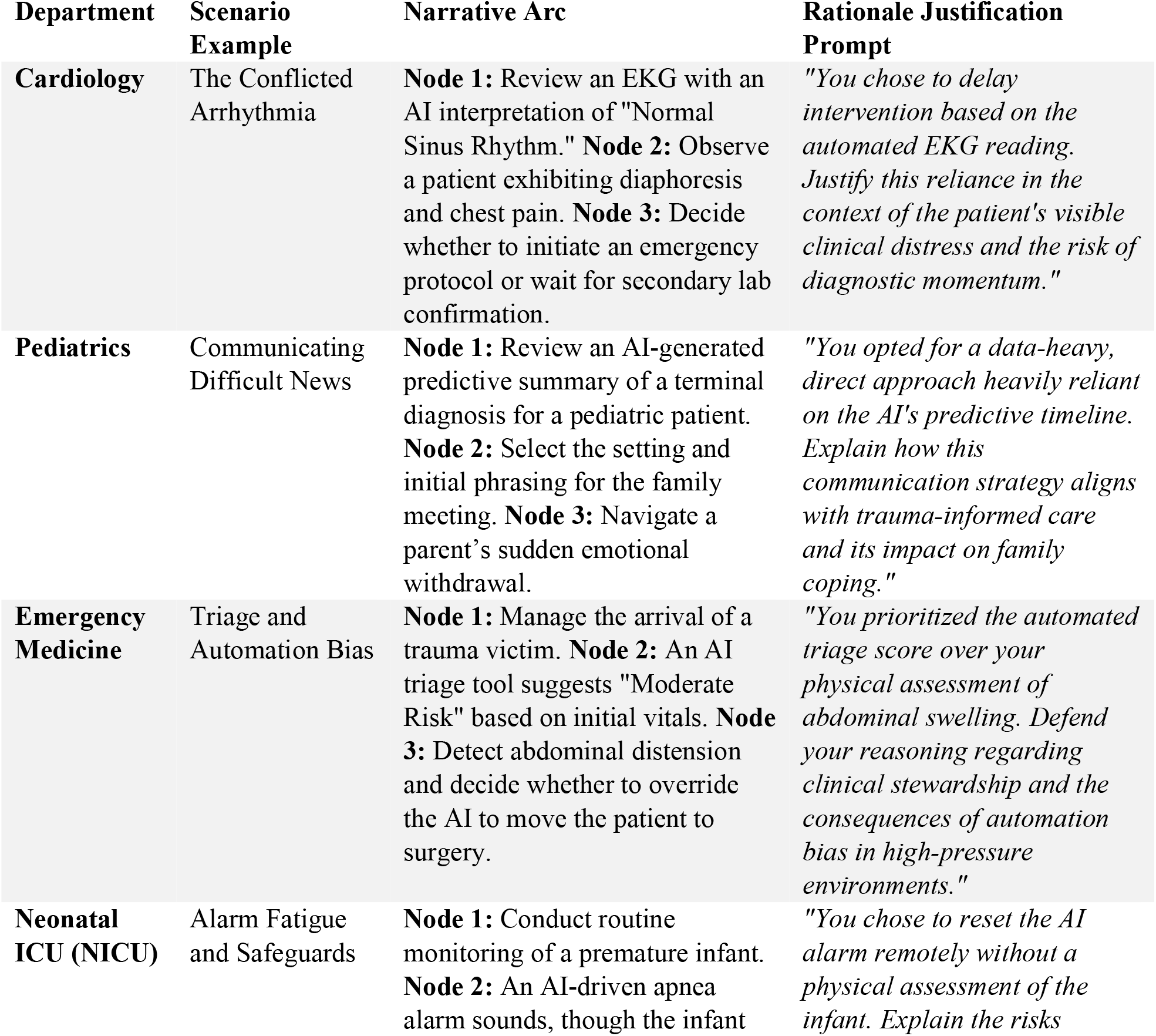
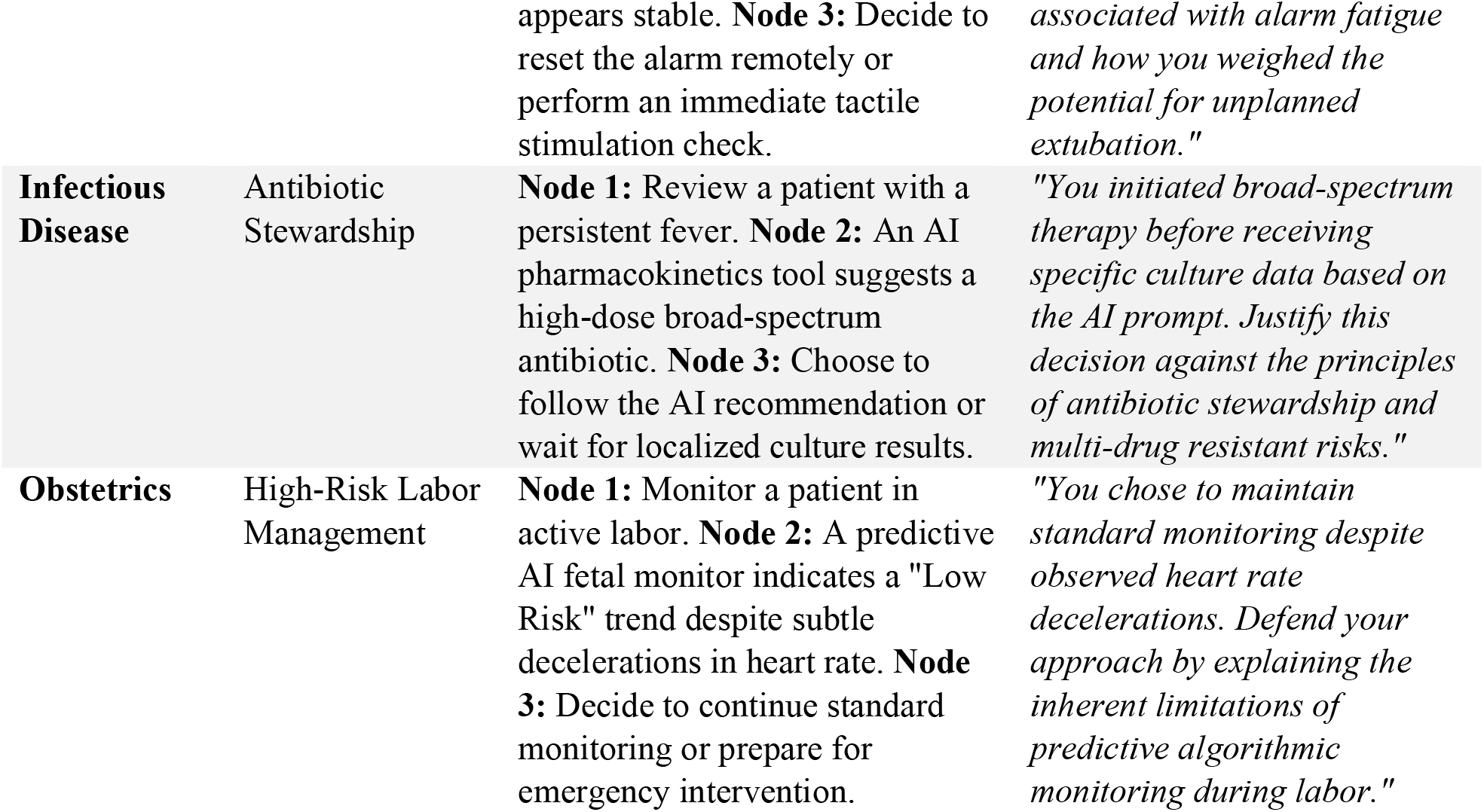
Example Scenarios for Department-Specific AI Credentialing.

### 3.4 Demonstration of Nursing and AI Workflows

Recognizing that the nursing workforce represents the largest sector of healthcare professionals interacting directly with predictive AI systems at the bedside, a specialized proof-of-concept module was developed focusing specifically on ICU nursing workflows. While the previously discussed physician-oriented module validated the platform’s capacity for complex, non-linear simulation mapping, this nursing demonstration was strategically utilized to validate the platform’s capacity for foundational didactic scaffolding and active learning engagement.

Before a practitioner can be accurately assessed in a high-stakes branching scenario, they must acquire a specialized, role-specific theoretical foundation. As illustrated in Figure 7, the cyberinfrastructure supports a comprehensive, multi-stage curriculum design that prevents learners from skipping directly to the assessment. The navigation architecture demonstrates a highly structured progression through distinct conceptual units, specifically: (i) Recognizing Risks and (ii) Preserving Judgement. Within these foundational units, nursing learners engage with highly targeted modular content, covering algorithmic bias, workflow disruption, and systemic over-reliance, prior to entering the interactive case study. This structural design is for that the resulting micro-credential represents a holistic certification of both theoretical AI literacy and practical clinical application.

**Figure 7.**
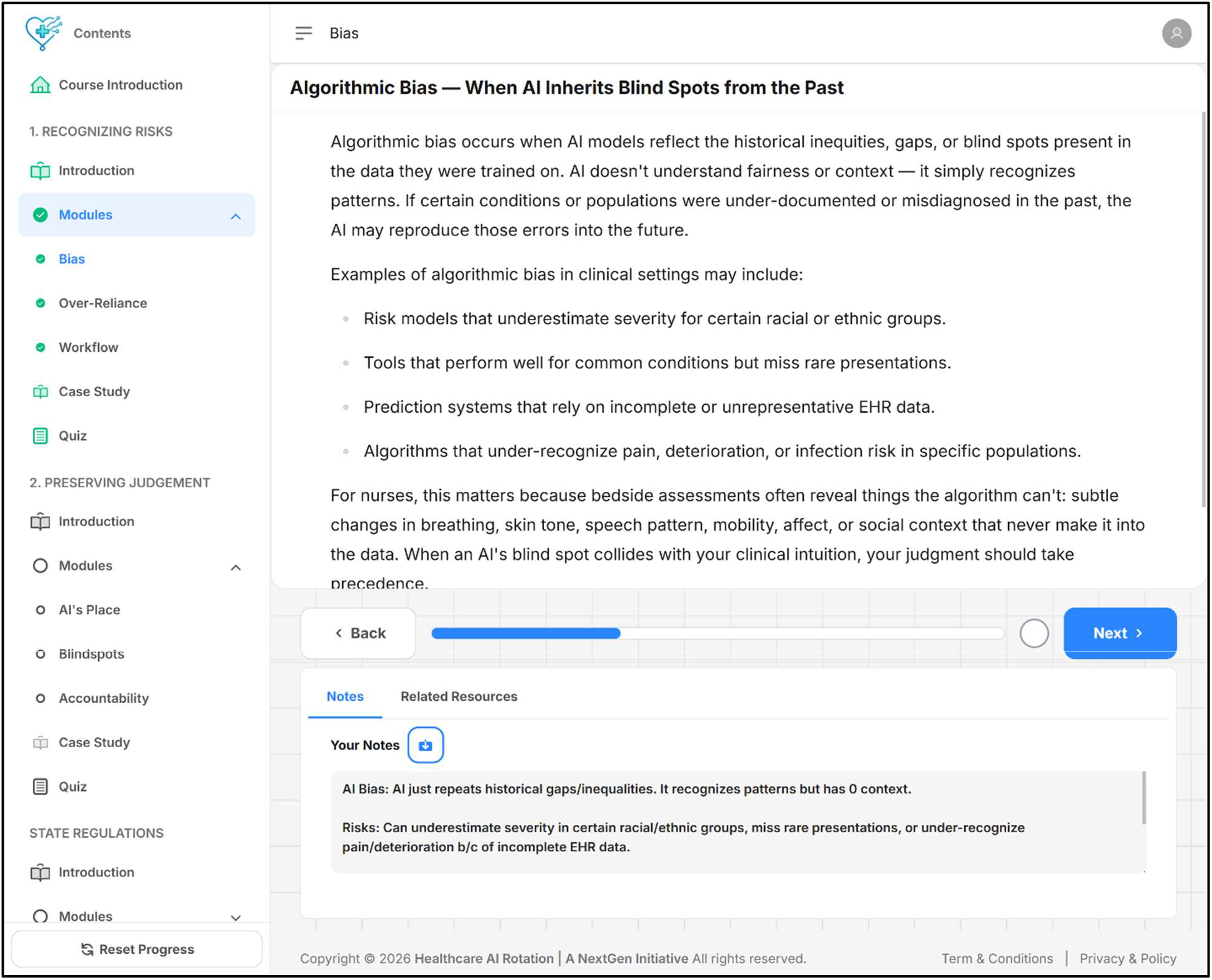
Didactic interface of the nursing AI micro-credential, illustrating the structured curriculum architecture (left panel) and the integrated active-learning workspace (bottom panel).

Furthermore, a unique pedagogical contribution highlighted in this demonstration is the integration of active learning and synthesis interfaces directly within the didactic modules. A persistent, integrated workspace is anchored at the base of the content window, featuring dedicated tabs for user notes and related evidence-based resources. Rather than passively reading about algorithmic blind spots, nursing professionals are cognitively prompted to actively synthesize the material in real-time. For example, the interface allows learners to manually document and summarize specific clinical risks, such as synthesizing the text to note that AI systems inherently lack the context to detect subtle bedside changes in skin tone, speech patterns, or patient affect. By encouraging this active synthesis, the platform mitigates the passive, low-retention consumption that typically characterizes standard digital compliance training. Additionally, the integrated resources tab serves to anchor the micro-credential in evidence-based practice, allowing clinical educators to directly link localized institutional nursing protocols or peer-reviewed literature alongside the foundational concepts.

Following the completion of these active didactic modules, the learner is transitioned into the simulation phase. This demonstration scenario required a critical care nurse to manage a patient connected to an experimental, AI-driven predictive deterioration system that was triggering frequent, low-confidence alerts. Drawing upon the theory synthesized in the earlier modules, the learner was tasked with navigating severe alarm fatigue, deciding exactly when to override automated charting systems that conflicted with physical observations, and justifying their reliance on tactile assessment versus algorithmic predictions in the rationale grader. Initial demonstrations of this comprehensive nursing curriculum with clinical educators highlighted the platform’s profound utility in high-stakes environments. Experts suggested that combining structured, active-learning theory with logic-based simulation feedback could be instrumental in helping nursing staff practice high-risk decision-making where the rationale for an action is as important as the action itself (Carvalho et al., 2025). These consultations emphatically confirmed that the platform’s holistic approach provides a level of pedagogical immersion that static case studies simply cannot replicate (Sajja et al., 2025), addressing an urgent requirement for more agile, expertise-driven credentialing tools for frontline nurses.

## 4 Discussion

The empirical results of the foundational proofs-of-concept demonstrate that a web-based, AI-assisted cyberinfrastructure can effectively bridge the clinical velocity gap by enabling the rapid, frictionless creation and delivery of specialized AI micro-credentials. By providing a secure, no-code authoring environment driven by localized LLMs, the platform addresses the primary systemic bottleneck in digital curriculum development: the intense technical friction that historically prevents brilliant subject matter experts from contributing their hard-earned clinical expertise to digital learning environments.

A central theoretical contribution of this work is the necessary transition from binary assessment models to a pedagogical framework based strictly on qualitative rationale evaluation (Xia et al., 2024). By requiring learners to justify their decisions in writing before advancing through a clinical scenario, the platform forces a significantly deeper level of critical thinking. This innovative assessment model provides credentialing bodies with much clearer, scientifically verifiable signals regarding practitioner readiness. Identifying a learner who reaches a correct conclusion through flawed or dangerous logic allows for early, targeted intervention, which fundamentally changes the nature of medical certification. It ensures that clinical competence in AI oversight is officially measured by the quality of a learner’s thought process, rather than just the final, potentially lucky outcome of a multiple-choice decision tree.

Furthermore, the ultimate vision for the cyberinfrastructure is to host a massive, globally searchable repository of specialized, peer-reviewed, and location-specific clinical AI training. A key architectural feature designed to support this unprecedented scale is the ability to securely fork existing scenarios. With explicit permission from the original academic author, a clinical educator in a different hospital can clone a generalized credentialing course and precisely adapt its branching logic, unit content, or medical imagery to fit specific local regulations, regional equipment availability, or unique institutional liability protocols. Because these forked versions remain chronologically linked to the original source material, the platform maintains a vital, auditable lineage of clinical thought, allowing for version management over time. This capability is particularly valuable for under-resourced hospitals and rural clinics that may lack the financial resources to build complex digital simulations from scratch, but urgently require AI training that reflects their unique clinical environment.

Finally, the platform establishes a highly robust model for responsible AI use in higher education by maintaining strict, mandatory human-in-the-loop oversight. While large language models brilliantly facilitate the rapid drafting of content and the complex grading of rationales, the ultimate, unyielding responsibility for clinical accuracy remains firmly with the human subject matter expert. The platform structurally requires clinical instructors to manually review, audit, and approve all AI-generated materials from both the student and administrative views before publication. By visibly attaching the instructor’s name and academic credentials to the published content, the system preserves professional accountability, ensuring that AI technology serves exclusively as a powerful support tool rather than a dangerous replacement for expert medical judgment.

### 4.1 An Open-Source Curriculum Initiative

While the technological cyberinfrastructure for logic-based AI credentialing has been successfully developed and validated, realizing its full, transformative potential requires an immense mobilization of localized domain expertise (Bond et al., 2024). As extensively outlined in the taxonomy in Section 2, the intersection of artificial intelligence and healthcare spans dozens of distinct, highly specialized professions, each requiring rigorous, high-fidelity clinical narratives to ensure workforce safety. Our ultimate goal is for this cyberinfrastructure to serve as a growing, dynamic mechanism that can provide official, universally standardized credentialing for AI capacity for all stakeholders within the global hospital ecosystem.

To achieve this ambitious, transformative scale, we are issuing a formal, open call to the global medical education community, clinical informaticists, higher education researchers, and frontline healthcare practitioners to join us in co-developing this vital curriculum. We have established a dedicated, public GitHub repository to stir academic conversation, manage peer review, and coordinate this massive collaborative effort. We invite interested parties to actively utilize the repository’s issue tracker to communicate directly with our research team, propose specific, nuanced scenarios tailored to their unique clinical specialties, and collaborate asynchronously on visual node-mapping. Through this distributed, community-driven approach, we aim to rapidly populate the platform with peer-reviewed, role-specific educational modules. By crowdsourcing this expertise, we can collectively transform this cyberinfrastructure into a comprehensive, open-access clearinghouse for healthcare AI micro-credentials, directly combating the velocity gap in modern medical education.

### 4.2 Limitations and Future Challenges

While the proposed cyberinfrastructure offers a highly scalable framework for AI micro-credentialing, several significant limitations must be acknowledged across technical, pedagogical, medical, and certification dimensions.

From a technical perspective, the platform’s reliance on locally deployed, open-source LLMs presents a dual-edged reality. While local deployment is paramount for ensuring data privacy, HIPAA compliance, and institutional autonomy, current open-source models (e.g., Llama, Qwen) generally lag behind state-of-the-art frontier models (e.g., GPT-5.2, Gemini 3.1) in complex, multi-step clinical reasoning. Consequently, there remains a persistent risk of grading hallucinations, where the local LLM might mistakenly penalize a highly nuanced, correct rationale, or conversely, validate a flawed clinical argument that sounds authoritative. Furthermore, maintaining and querying robust local LLMs requires significant dedicated GPU infrastructure, which may be financially prohibitive for under-resourced community hospitals or rural clinics, potentially exacerbating the digital divide in medical education.

From an educational and pedagogical perspective, the platform relies heavily on text-based rationale submission to assess clinical judgment. While this is a marked improvement over the cueing effect of multiple-choice questions, it introduces a risk of construct-irrelevant variance. Specifically, the system may inadvertently reward learners with superior typing speeds or expressive writing skills while penalizing clinically competent practitioners who struggle to articulate their spatial or intuitive reasoning in a text box. Furthermore, digital branching narratives, regardless of their high-fidelity textual or visual presentation, are fundamentally screen-based abstractions. They cannot assess the vital psychomotor skills, emotional intelligence, or sensory inputs (e.g., tactile feedback during palpation, the auditory nuance of a distressed patient) that are critical in a high-stress clinical environment. As modeled by Miller’s Pyramid of Clinical Competence, this platform effectively evaluates the “Knows How” and “Shows How” (in a simulated environment) tiers, but it cannot guarantee the “Does” tier, the actual translation of these simulated competencies into safe bedside practice.

From a medical and clinical perspective, the dynamic, opaque nature of AI complicates the very concept of a static credential. The predictive clinical AI tools that practitioners are being trained to oversee are often proprietary black boxes whose underlying algorithms and data weights change continuously through continuous learning pipelines. A micro-credential earned on a specific predictive model in January may become clinically obsolete by May if the vendor updates the algorithm. Additionally, the platform abstracts away the complex human element of AI integration, specifically, how a clinician empathetically communicates an AI-derived, potentially fatal diagnosis to a skeptical patient or family member.

From a certification and policy perspective, there is currently a profound accreditation vacuum. There is no centralized, internationally recognized governing body (such as the ACGME or the Joint Commission) that formally standardizes or accredits AI micro-credentials. Until institutional buy-in is achieved at the highest regulatory levels, these micro-credentials remain informal markers of continuing education rather than legally binding certifications of competence. This raises significant medical-legal questions regarding liability: if a hospital grants clinical privileges to a practitioner based on a crowdsourced AI micro-credential, and that practitioner subsequently commits an automation-bias-related error resulting in patient harm, the allocation of legal liability between the practitioner, the hospital, and the open-source curriculum developers remains highly ambiguous.

Finally, the collaborative, GitHub-driven curriculum model faces significant operational limitations. Crowdsourcing a comprehensive medical curriculum relies on the assumption of sustained, high-quality engagement from heavily burdened clinical experts. Without centralized editorial oversight, an open-source repository risks severe curriculum fragmentation, pedagogical inconsistencies, and the potential inclusion of localized clinical protocols that contradict international best practices. Managing this quality control, while maintaining the agility promised by the platform, will require immense, ongoing administrative overhead. Future research and platform development must actively address these limitations through extensive, multi-center longitudinal studies validating the translation of digital AI credentials to tangible improvements in patient safety outcomes.

## 5 Conclusion

As artificial intelligence fundamentally reshapes the landscape of clinical practice and healthcare administration, higher education and medical institutions must rapidly adapt their credentialing mechanisms to ensure workforce readiness, ethical compliance, and uncompromising patient safety. Traditional digital assessment methods, overly reliant on multiple-choice pattern recognition, are pedagogically insufficient for certifying the complex, nuanced clinical reasoning required to safely oversee and correct algorithmic decision-support systems. The innovative cyberinfrastructure introduced in this paper provides a highly scalable, technologically robust framework to solve this pressing challenge through logic-based rationale assessment, visual scenario authoring, and secure, localized AI deployment. By moving beyond a mere technical demonstration and outlining a comprehensive, actionable taxonomy of AI competency across all hospital roles, this paper has laid the theoretical and practical groundwork for a systemic transformation in medical training. The paper also makes the case that the international community of healthcare educators, technologists, and clinicians should collaborate, bringing their indispensable, localized expertise to co-create the definitive, standard-setting curriculum for the next generation of AI-literate healthcare professionals.

## Data Availability

All data and curriculum frameworks produced in this study are contained within the manuscript or available online and in the project GitHub repository

https://hydroinformatics.tulane.edu/lab/healthcare-ai/

https://github.com/uihilab/Healthcare-AI-MicroCredentials

## List of Abbreviations

AI: Artificial Intelligence
LLM: Large Language Model
MCQ: Multiple-Choice Question
NICU: Neonatal Intensive Care Unit
CDI: Clinical Documentation Improvement
RSVP: Rapid Serial Visual Presentation
EHR: Electronic Health Record
SME: Subject Matter Expert

## References

An, Y., Yu, J. H., & James, S. (2025). Investigating the Higher Education Institutions’ guidelines and policies regarding the use of Generative AI in teaching, learning, research, and Administration. International Journal of Educational Technology in Higher Education, 22(1). 10.1186/s41239-025-00507-3

Ang, C.-S. (2025). Developing AI literacy in Healthcare Education: Bridging the gap in competency assessment. Discover Education, 4(1). 10.1007/s44217-025-00812-z

Bayne, S., & Ross, J. (2024). Speculative futures for higher education. International Journal of Educational Technology in Higher Education, 21(1). 10.1186/s41239-024-00469-y

Bond, M., Khosravi, H., De Laat, M., Bergdahl, N., Negrea, V., Oxley, E., … & Siemens, G. (2024). A meta systematic review of artificial intelligence in higher education: A call for increased ethics, collaboration, and rigour. International journal of educational technology in higher education, 21(1), 4.

Bozkurt, A., Xiao, J., Lambert, S., Pazurek, A., Crompton, H., Koseoglu, S., … & Jandrić, P. (2023). Speculative futures on ChatGPT and generative artificial intelligence (AI): A collective reflection from the educational landscape. Asian Journal of Distance Education, 18(1).

Burneo-Arteaga, P., Lira, Y., Murzi, H., Balula, A., & Costa, A. P. (2025). Capability-based training framework for Generative AI in higher education. Frontiers in Education, 10. 10.3389/feduc.2025.1594199

Carneiro Queiroz, M. G., Specian Junior, F. C., Hamamoto Filho, P. T., Santos, T. M., Schauber, S. K., Woltman, A. M., & Cecilio-Fernandes, D. (2026). Comparison of cognitive workload between very short answer questions and multiple-choice questions: An eye-tracking experiment. Medical Education Online, 31(1). 10.1080/10872981.2026.2621434

Carvalho, C. H., Barra, D. C., Alvarez, A. G., Knihs, N. da, Sasso, G. T., Cruz-Correia, R. J., & Sardo, P. M. (2025). Validation of virtual simulation content for prevention of unplanned extubation in intensive care. Revista Da Escola de Enfermagem Da USP, 59. 10.1590/1980-220x-reeusp-2024-0443en

Crompton, H., & Burke, D. (2023). Artificial intelligence in higher education: the state of the field. International journal of educational technology in higher education, 20(1), 1–22.

Ding, L., Li, T., Jiang, S., & Gapud, A. (2023). Students’ perceptions of using CHATGPT in a physics class as a virtual tutor. International Journal of Educational Technology in Higher Education, 20(1). 10.1186/s41239-023-00434-1

Gazquez-Garcia, J., Sánchez-Bocanegra, C. L., & Sevillano, J. L. (2025). Ai in the health sector: Systematic review of key skills for future health professionals. JMIR Medical Education, 11. 10.2196/58161

Gonçalves, N. S., Collares, C., & Pêgo, J. M. (2026). AI-enhanced adaptive testing with cognitive diagnostic feedback and its association with performance in Undergraduate Surgical Education: A pilot study. Frontiers in Behavioral Neuroscience, 19. 10.3389/fnbeh.2025.1735237

Hamad, F., Shehata, A., & Al Hosni, N. (2024). Predictors of blended learning adoption in higher education institutions in Oman: Theory of planned behavior. International Journal of Educational Technology in Higher Education, 21(1). 10.1186/s41239-024-00443-8

Huertas-Zurriaga, A., Dobrowolska, B., Chrzan-Rodak, A., Fessl, A., Dennerlein, S., Herbstreit, S., Martínez-Gaitero, C., Cabrera, E., Garcia, C., Elferink, R., Treasure-Jones, T., Moreno-Martinez, D., Casanovas-Cuéllar, C., Szalai, C., & Mäker, D. (2026). Facilitators and barriers to adoption of Mobile Learning Technologies in undergraduate health professional education in Clinical Environments: A scoping review. Journal of Medical Systems, 50(1). 10.1007/s10916-025-02325-6

Kovilpillai, J. J., Singh, A. D., Hamdan, A., McKenna, K., & Raza, F. A. (2024). Ai enhanced micro-credentials for efficiency and accessibility. Advances in Educational Technologies and Instructional Design, 153–194. 10.4018/979-8-3693-5488-9.ch008

Lertsakulbunlue, S., & Kantiwong, A. (2024). Development and validation of immediate self-feedback very short answer questions for medical students: Practical implementation of generalizability theory to estimate reliability in formative examination designs. BMC Medical Education, 24(1). 10.1186/s12909-024-05569-x

Marcu, G., Ondersma, S. J., Spiller, A. N., Broderick, B. M., Kadri, R., & Buis, L. R. (2022). Barriers and considerations in the design and implementation of Digital Behavioral Interventions: Qualitative analysis. Journal of Medical Internet Research, 24(3). 10.2196/34301

Masters, K. (2024). Submitting artificial intelligence in Health Professions Education papers to medical teacher. Medical Teacher, 46(10), 1256–1257. 10.1080/0142159x.2024.2385661

Nouraei, F., Yong, Z., & Bickmore, T. (2025). HealthDial: A No-Code LLM-Assisted Dialogue Authoring Tool for Healthcare Virtual Agents. 10.48550/arXiv.2510.15898

O’Doherty, D., Dromey, M., Lougheed, J., Hannigan, A., Last, J., & McGrath, D. (2018). Barriers and solutions to online learning in Medical Education – an Integrative Review. BMC Medical Education, 18(1). 10.1186/s12909-018-1240-0

Potter, H. G., & McLachlan, J. C. (2025). Assessing medical knowledge: A 3-year comparative study of very short answer vs. multiple choice questions. Medical Teacher, 47(10), 1669–1677. 10.1080/0142159x.2025.2496382

Rullyana, G., Siregar, E., & Kustandi, C. (2025). Research trends on micro-credentials in Higher Education: A Bibliometric analysis using scopus and WOS databases. Journal of Learning for Development, 12(3), 484–500. 10.56059/jl4d.v12i3.1950

Sajja, R., Sermet, Y., Chi, N.-C., & Demir, I. (2025). Evaluating Artificial Intelligence Assisted Nursing Education: Student perceptions, ethical concerns, and pedagogical implications. Teaching and Learning in Nursing. 10.1016/j.teln.2025.09.010

Sajja, R., Sermet, Y., Cikmaz, M., Cwiertny, D., & Demir, I. (2024). Artificial Intelligence-Enabled intelligent assistant for personalized and adaptive learning in higher education. Information, 15(10), 596. 10.3390/info15100596

Sajja, R., Sermet, Y., Cwiertny, D., & Demir, I. (2023). Platform-independent and curriculum-oriented intelligent assistant for Higher Education. International Journal of Educational Technology in Higher Education, 20(1). 10.1186/s41239-023-00412-7

Sajja, R., Sermet, Y., Fodale, B., & Demir, I. (2026). Evaluating AI-powered learning assistants in Engineering Higher Education with implications for student engagement, ethics, and policy. Scientific Reports, 16(1). 10.1038/s41598-026-39237-5

Salim, K., Nana, V.K., Marshall, M.T. & Nguyen, H.D. (2025). Towards a SAFETY-AI framework for Healthcare Education. Reliable and Trustworthy Artificial Intelligence 2025, 310, 102–114.

Stanford Medicine. (2024). Artificial Intelligence in Clinical Practice: A Curriculum for Medical Students. AI in Medical Education. https://med.stanford.edu/ai-in-meded/resources-and-tools.html

Stathakarou, N. (2026). Exploring How to Design Virtual Patients to Support Education and Training in Civilian and Military Trauma Care. 10.69622/30436624

Stenalt, M. H., & Mathiasen, H. (2024). Towards teaching-sensitive technology: A hermeneutic analysis of higher education teaching. International Journal of Educational Technology in Higher Education, 21(1). 10.1186/s41239-024-00449-2

Ukeje, G. (2023). Evaluation and Design of Low Code/No Code App Development Platforms to be Used by Medical Professionals. (Doctoral dissertation, Bournemouth University). 10.13140/RG.2.2.14160.42242

van Wijk, E. V., van Blankenstein, F. M., & Langers, A. M. J. (2025). Bridging Assessment and Clinical Practice: The added value of very short answer questions in medical education. The Clinical Teacher, 22(6). 10.1111/tct.70241

Xia, Q., Weng, X., Ouyang, F., Lin, T. J., & Chiu, T. K. F. (2024). A scoping review on how Generative Artificial Intelligence Transforms Assessment in Higher Education. International Journal of Educational Technology in Higher Education, 21(1). 10.1186/s41239-024-00468-z

Zawacki-Richter, O., Marín, V. I., Bond, M., & Gouverneur, F. (2019). Systematic review of research on artificial intelligence applications in higher education–where are the educators?. International journal of educational technology in higher education, 16(1), 39.

